# Instagram as a health education tool: Evaluating the efficacy and quality of medical content on Instagram in Azerbaijan

**DOI:** 10.1101/2023.12.03.23299316

**Authors:** Bahar Graefen, Shams Hasanli, Araz Jabrayilov, Gulgaz Alakbarova, Khayala Tahmazi, Jamila Gurbanova, Nadeem Fazal

## Abstract

**Background:** In recent years, Instagram has become the most popular tool among professional doctors in Azerbaijan for educating their patients. The use of the Instagram application aims to raise patients’ awareness of the importance of taking care of their health and to increase their knowledge about their health conditions using modern services. In this article, the authors examine the quality of Instagram content for health education among the population.

**Methods:** We conducted a survey to collect anonymous data from more than 205 respondents and summarized the following points.

**Results:** 65% of the respondents were already obtaining health information from Instagram before to participating in the study. 15.1 % of them frequently visit Instagram for health information while 5% had found the health information accessed there harmful. 71% of respondents think accessing health information in this way is beneficial but that the quality and usefulness of the content is average. 95% of respondents reported that the health information they obtained from the identical platform was not causing them any harm

**Conclusion:** The medical information shared on Instagram is generally considered useful and beneficial by the population, but it is desirable to improve the quality of the content.

## INTRODUCTION

Today, the Internet is a public, collaborative, and self-sustaining facility accessible to millions of people globally. It is currently used by many people as a primary source of information. It has also enabled the creation of its own social ecosystem through Internet browsers, search engines, social media and content sharing platforms. Examples of these Internet browsers include Mozilla Firefox®, Internet Explorer® and Google Chrome® (1). The popularity of social media has increased greatly in the last few years. Social media can be defined as “a group of internet-based applications that enable the creation and sharing of user-generated content”. Social media and content sharing platforms include Instagram®, Twitter®, Facebook®, YouTube®, TikTok®, WhatsApp®, etc. (2). The boom in technology has resulted in an increase in the number of materials and resources available to people (3). A number of studies show that social media can be a valuable resource for information exchange and collaboration (4). The development of social networks in rapidly developing countries such as Azerbaijan is closely linked to the rapid penetration of elements of the digital economy. For this reason, social media is of particular importance as a tool for education, business and social communication (5).

As of 2022, Azerbaijan’s population is 10 156 464, of which 52.9% is urban and 47.1% is rural (6). According to statistical research, social media, which is widely used around the world, has 4.15 million users in Azerbaijan. This corresponds to approximately 40% of the country’s population (7). Social media platforms represent some of the most beneficial marketing tools in Azerbaijan, offering the required value and focus to advertising, healthcare, and educational institutions. These social media channels have various distinct features and characteristics. Educational institutions can build direct connections with customers, get knowledge of their interests and preferences, and enhance their services by utilizing these platforms, which facilitate the sharing of numerous videos and pictures. This makes it simpler and more comfortable for businesses to interact with users more effectively (8). In addition, Instagram, which is the most frequently used social media network today and first appeared in 2010, is gaining growing popularity around the world as a photo sharing platform, and over time, new features such as video, messaging, and story sharing have been added to it (9). Instagram is a social networking website where users may share photos and videos. It is owned by Meta Platforms, an American corporation (10). Customers can utilize the app to send material throughout channels, which can be coordinated and exchanged by geotagging and hashtags. Currently, there are 1.35 billion monthly active users of Instagram, making up 17.6% of the total population. According to statistics, in 2021, Instagram users used the platform for an average of 30 minutes a day (11).

Instagram is fundamentally designed for entertainment and social connection. However, as it began to be seen as a tool that could also be used for educational purposes, a specific usage of Instagram emerged that had not been foreseen. Instagram’s enormous popularity and attractive interface present a unique chance to explore the possibilities of such platforms as non-traditional learning tools. Numerous novel ideas have emerged as a result of the usage of Instagram in educational settings, and these ideas now need more research. Creating a specific Instagram account for an educational setting and sharing details about assignments, projects, and events on a regular basis promotes transparency and informs people. Instagram users also utilize its image and video editing features to create educational posts (12). Health information, which is one of the leading content areas for social media platforms that contain various educational topics, is among the common reasons for applying to these platforms today (13).

As patients easily find information about their health issues online, they need fewer doctor’s recommendations (14). Therefore, the Internet has become an important source of information about healthcare worldwide. It was estimated that 84% of individuals over the age of 18 frequently interacted with the Internet in 2015. This corresponds to 6.165 billion people. Additionally, social media and video sharing platforms such as Instagram®, Twitter®, YouTube®, and Facebook® as well as other search engines such as Google®, Bing®, Yahoo® etc. are also popular sources of health information. For example, more than 2 billion people visit video sharing platforms such as Instagram® and YouTube® every month. This is because, among other reasons, they host a rich collection of health education videos. (15)

Many individuals often utilize social media in health-related situations (16). 80 percent of cancer patients use social media as a personal tool to interact with other patients (17). Over 80% of state health departments in the United States (US) have social media user profiles inside healthcare organizations (18). Among healthcare professionals, 65% of radiologists in the USA and Europe use social media for various health-related reasons (19). A review of current technology can provide guidance to practitioners considering the use of social media and to researchers seeking to advance our understanding of the use of social media for health purposes.

Specifically, social media is increasingly becoming a supportive tool in healthcare by allowing its users to obtain and share information. It allows healthcare professionals to connect with others in the field and communicate with colleagues, patients or the public regarding health issues. Additionally, social media supports patient empowerment by expanding patients’ knowledge and placing them in a position where they can take control of their own health needs (20). After educating themselves online before going to the doctor, patients can apply to the hospital with a fixed diagnosis in mind and a preferred treatment option based on this information. This may affect their management decisions (21, 22). However, it should be taken into consideration that content published on social media platforms may have disadvantages as well as advantages.

It is generally accepted that using modern information and communications technologies in healthcare has several benefits (23). Social media offers an immense amount of potential advantage in this broad area of new developments in healthcare, according to the research. This is so because social media provides novel opportunities for information access and distribution, promotes collaboration and social support, and ensures the contribution of the relevant parties (24, 25).

In addition to these advantages, there are also some disadvantages of using social media as a tool in the health sector. These include verifying the accuracy of information, maintaining confidentiality under the Health Insurance Portability and Accountability Act of 1996 (HIPAA) regulations, and the lack of a robust peer review system (26,27). Although research has been conducted on the benefits, risks, and use of social media in health education, little is known about the specific use of social media to support health education and attitudes towards social media (28). In addition, considering that there are no quality control measures or peer review processes to ensure the accuracy of health information and videos shared on the Internet, patients or healthcare professionals may be exposed to imprecise or deceptive information. The accuracy of patient education in online search engines and video sharing platforms has been investigated in the context of different diseases and conditions. The results show that these online search engines and video sharing platforms often have insufficient educational content, which can lead to misinformation (29).

As a result, a novel kind of professionalism, known as “e-professionalism,” has developed. E-professionalism has been defined by Cain and Romanelli (30) as attitudes and actions that conform to the usual professionalism paradigms but are conveyed through digital media. Unprofessional materials posted by healthcare workers on social media can vary from showing information about patients to negative remarks about colleagues to profanity, and may be considered differently based on social background and context (31). Instagram content that is widely shared around the world and contains health education is also common in Azerbaijan. Evaluating these medical contents from different perspectives is important in terms of increasing the awareness of the general population and reducing the rate of misinformation. Therefore, this study aimed to evaluate the effectiveness and quality of medical content on Instagram in Azerbaijan.

## METHODS

### Study Design

The questionnaire was created to collect information from participants anonymously. It included both closed-ended questions involving Likert scales. This research aimed to evaluate the efficacy and quality of medical content on Instagram as a health education tool.

The survey consisted of a series of questions related to Instagram usage for health information, perceived benefits, and content preferences. Key data points obtained from the survey include:

- Have you received health information from Instagram before?
- How often do you visit Instagram for health information?
- Have you been harmed according to the medical information you received from Instagram?
- Does the medical information you get from Instagram benefit you?
- Rate the usefulness of the medical information shared on Instagram (1 - low, 5 - high).
- Rate the quality of medical information shared on Instagram (1 - low, 5 - high).

### Data Collection

The study lasted 3 months in total, and the data was collected through a structured survey questionnaire. The survey was designed to assess the usage patterns and perceptions of Instagram as a source of health-related information. Students received a request to take part in the study by email from the researchers. After two follow-up emails were sent at three-week intervals, the online survey was ended after a total of eight weeks. Students weren’t provided with a reward or incentive for taking part in the study. The respondents were given comprehensive instructions on how to complete the survey and were promised that the results would be treated confidentially. Collected data was encrypted and was not shared or sold to third parties.

### Participants

The study participants consisted of 205 students from various institutions in Azerbaijan. The selection of this sample aimed to represent a diverse cross-section of students in the country.

### Data Analysis

Data analysis involved the computation of descriptive statistics to summarize the survey responses. Percentages were used to quantify the proportion of respondents for each survey item. These results provide insights into the prevalence and impact of Instagram as a health education tool among students in Azerbaijan.

### Ethical approval

The research adhered to ethical guidelines, ensuring the anonymity and confidentiality of participants. Informed consent was obtained from all participants before data collection.

The Human Research Ethics Committee (Scientific Research Institute of Obstetrics and Gynecology) approved the study (Protocol 3-28-10/3-123/2023). Informed consent was obtained from all participants in this study.

### Statistical analysis

The statistical analysis was conducted using SPSS software version 20.0 (SPSS Inc., Chicago, IL, USA).

## RESULTS

The results of the survey show that 134 (65.4%) of the respondents were female, 69 (33.7%) were males, and 2 (0.98%) did not provide any answer on their gender. According to our results, all respondents had easy access to Instagram, and 69% of them are on the platform daily, 23% weekly, while 8% visit Instagram less often than that. Also, results demonstrate that significant number of participants had used Instagram for health information at least once. This illustrates the increasing relevance of social media platforms like Instagram in the sharing of health information, as well as the need to ensure the quality of such content posted on these platforms. 205 respondents confirmed that they had indeed accessed health information on Instagram. 72 (35 %) participants indicated that they have never sought health information from this platform (Figure 1).

**Figure 1.**
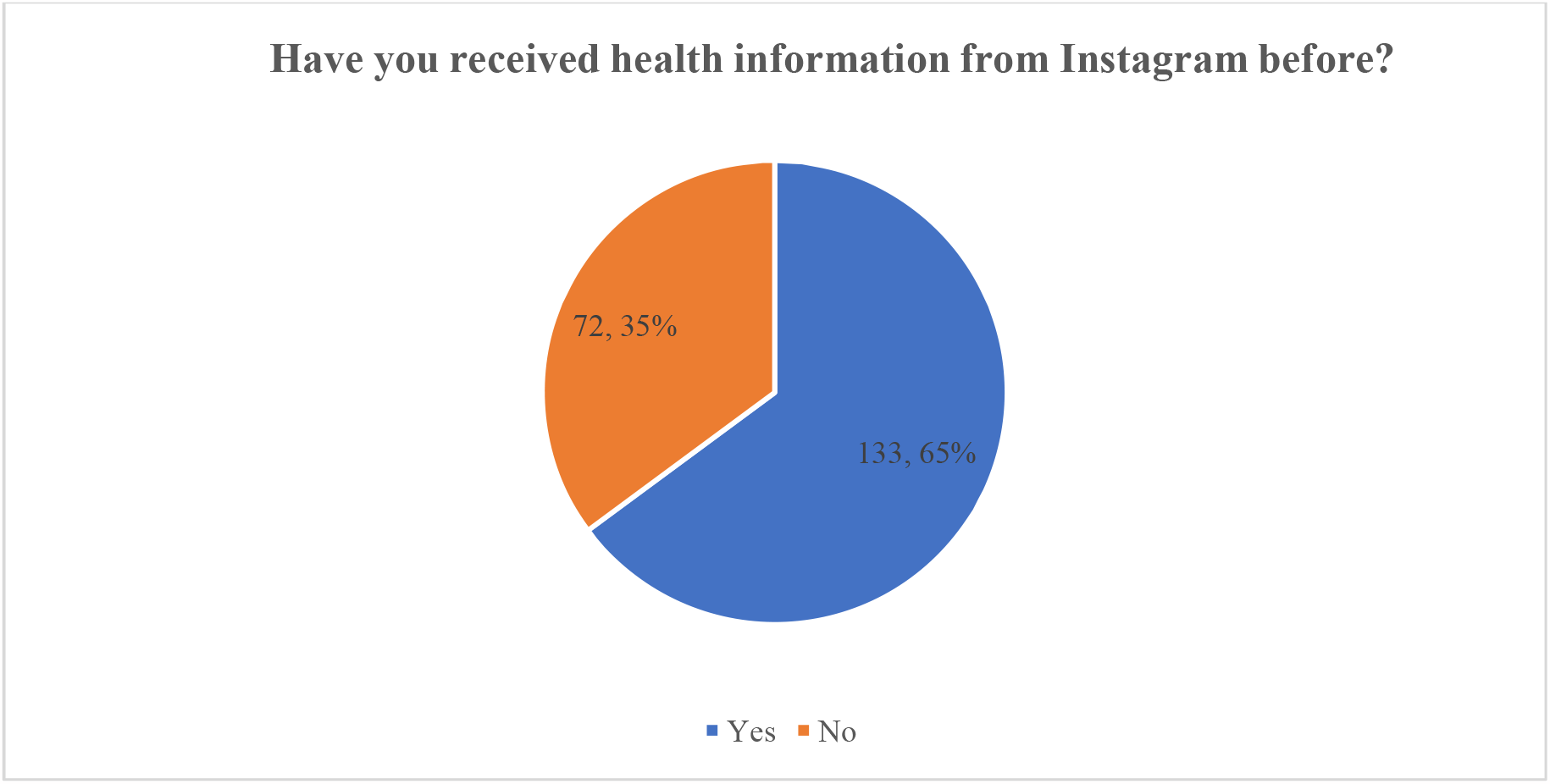
User engagement with health information on Instagram

As part of a research project studying the habits of individuals looking for health-related information on social media, especially Instagram, a survey was conducted to find out how frequently users use the platform for this reason in particular. The results indicate an extremely similar distribution among respondents who use Instagram never, rarely, or sometimes to obtain health information. Each category obtained 58 responses, or 28.3% of the total population asked for each group. It appears that although a significant segment of the population regularly utilizes the platform to access health-related material, there is a clear tendency towards rare rather than regular use. This may indicate that, despite its popularity, Instagram may not be the main source of health information for its users, or it could signal a skepticism regarding the validity of health-related content on social media (Figure 2).

**Figure 2.**
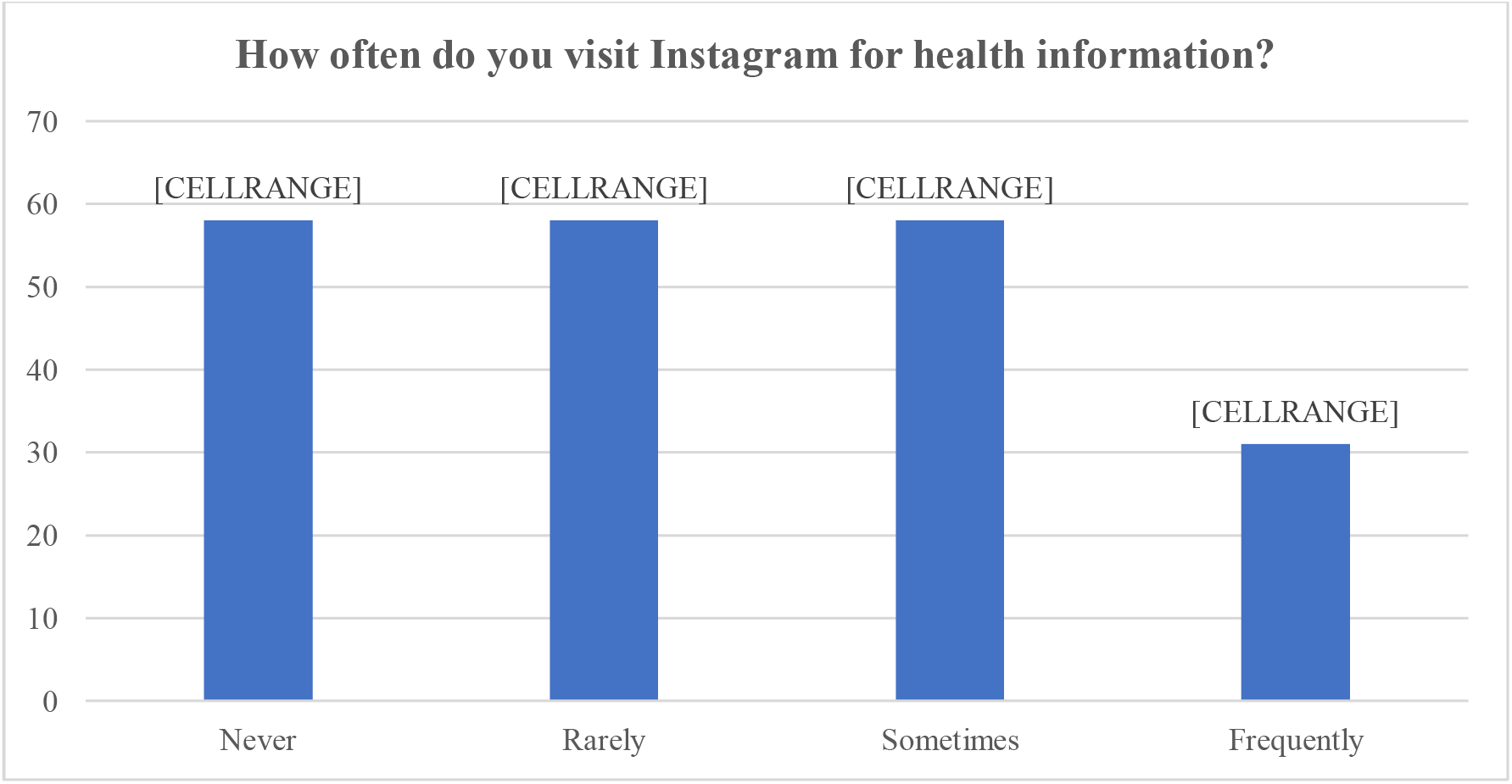
Frequency of accessing health information on Instagram

The results of the study demonstrated that 11 (5%) out of the total respondents, or a small percentage of the sample, confirmed that they had been harmed by medical information they had seen on Instagram. On the other hand, 192 (95%) respondents reported that the health information they obtained from the identical platform was not causing them any harm (Figure 3).

**Figure 3.**
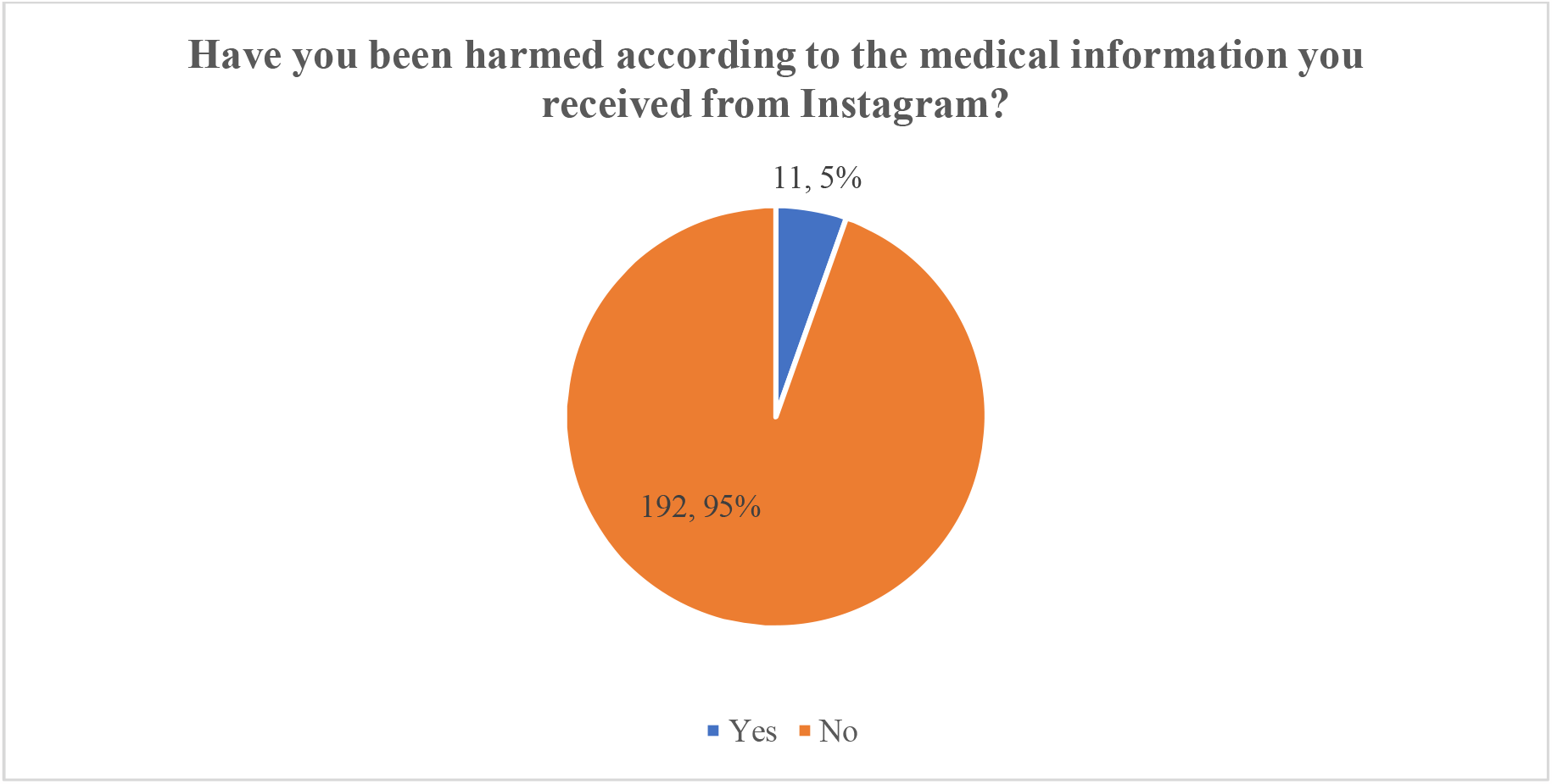
Impact of Instagram-sourced medical information on user harm

A significant number of respondents, 143 participants, stated in an online survey intended to evaluate the perceived advantages of the medical information acquired from Instagram that they found the contents to be beneficial. This reflects about 71 % of the group, suggesting that the vast majority of respondents considered the information provided to be valuable.

A recent study aimed at collecting user feedback focused on the perceived quality of medical content on Instagram. The majority of respondents, or around 65.2%, assessed the quality of medical information on Instagram as average, based on the data collected. It also suggests that although the content may be considered understandable and somewhat educational, its depth and authenticity may require some extra effort. Conversely, 20.2% of the participants reported that the quality of medical information was undesirable, which may be a sign of issues with the platform’s content’s source, accuracy, and usefulness. This view may be influenced by disinformation and the spreading of recommendations that are not supported by facts. On the other hand, just 14.6% of respondents thought the quality was outstanding (Figure 4).

**Figure 4.**
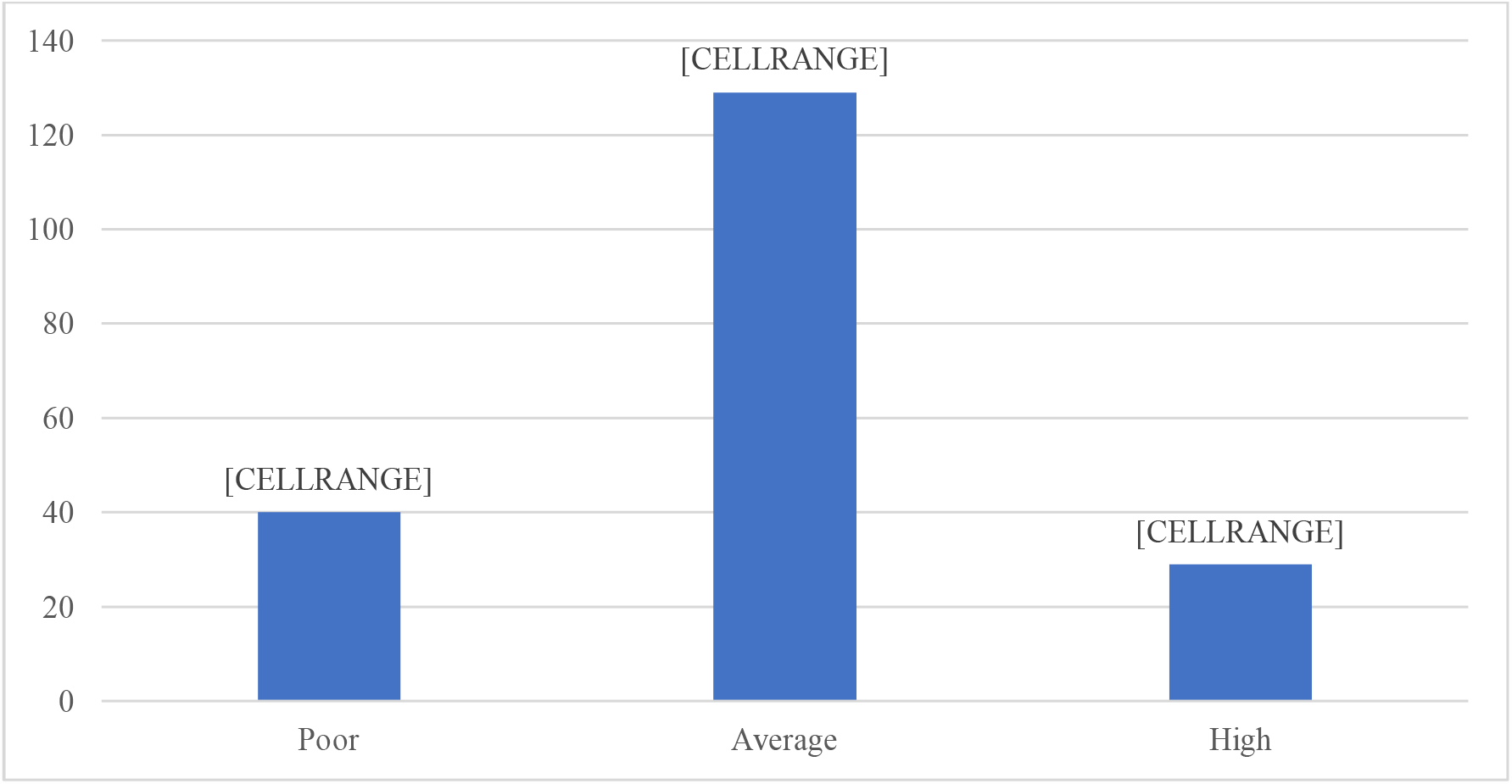
Rate the quality of medical information shared on Instagram

Participants were asked to rate the content’s perceived benefit on an Instagram scale ranging from 1 (low usefulness) to 5 (high usefulness). There is a range of viewpoints shown in the comments. A small percentage of the sample, 16.5%, had the lowest usefulness score of 1, indicating a significant lack of satisfaction with the usability of the medical information. 10% of participants assigned the content a usability rating of 2. Most answers ranged between the midpoint and the bottom of the scale, with 36% of participants giving the information’s utility a score of 3, which indicates a neutral viewpoint. Meanwhile, a significant number of the participants—24 %, to be exact—scored the utility as 4, indicating a positive perception of the value of the medical material. 13.5% of respondents rated the information with the highest usability rating of 5, indicating significant appreciation for it. (Figure 5).

**Figure 5.**
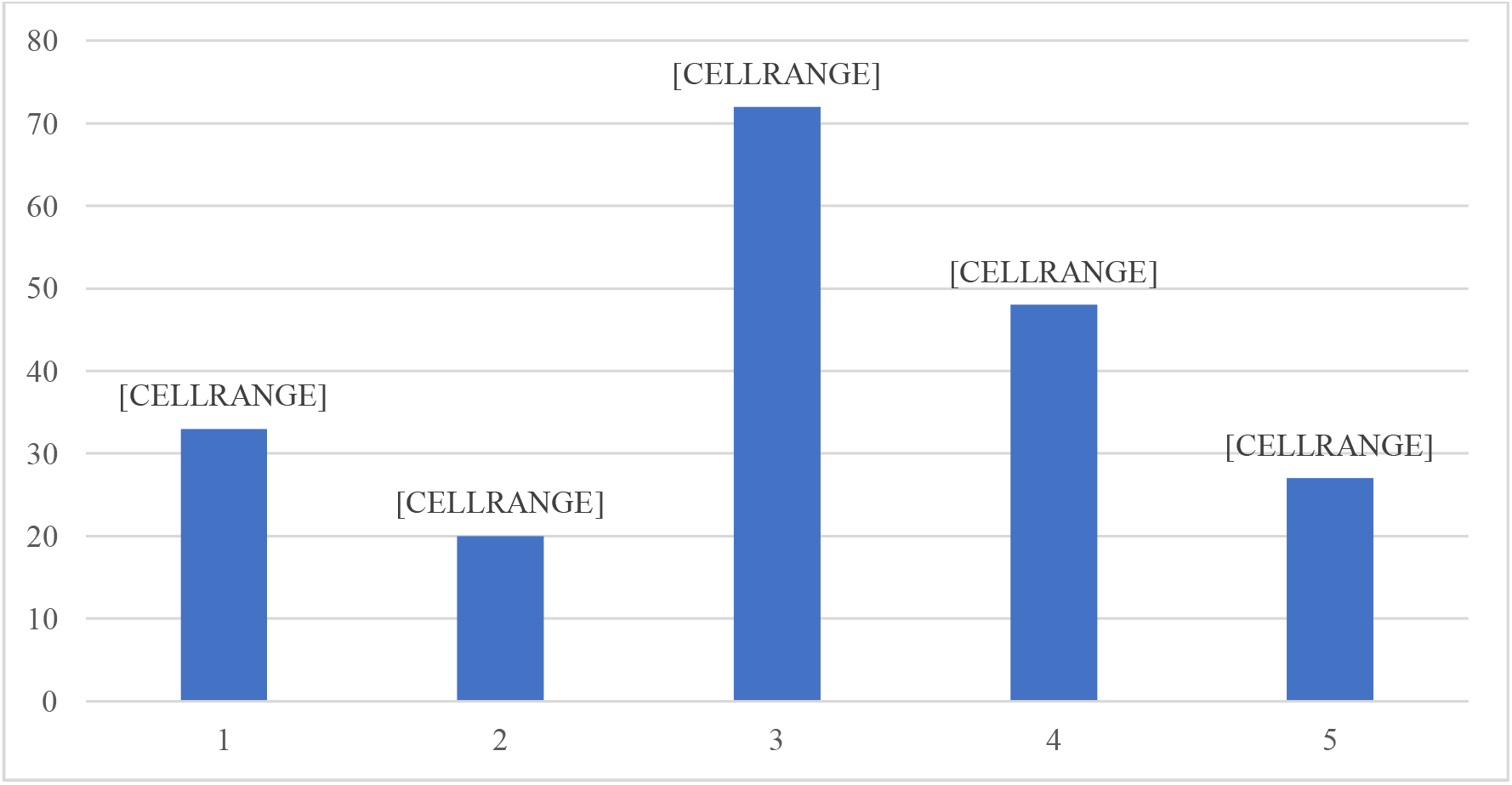
Rate the usefulness of the medical information shared on Instagram (1-low to 5 –high)

## DISCUSSION

The globally trend of Instagram’s contribution to health education distribution also exists in Azerbaijan, where the app’s broad popularity offers a unique opportunity to impact public health behaviors and knowledge. It is crucial to assess the medical components that is shared on these platforms because, in today’s digital age, knowledge spreads more quickly than ever. This is not only a question of academic interest; public health requires it. Health education can reach an extensive number of people in Azerbaijan and other countries because of the simplicity with which information can be shared on Instagram, which means that it can reach even people who might not otherwise have access to it. On the other hand, the ease of access also increases the possibility of disinformation disseminating rapidly compared to valid information. Public health problems may be worsened by misinformation, which can result in incorrect choices about an individual’s health and a general mistrust of reliable medical recommendations. Therefore, an assessment of medical information on Instagram in Azerbaijan is required to be detailed and extended. It should consider the information’s accuracy and reliability, the skills of the individuals presenting it, and how well it engages the public. It is not sufficient for the information to be accurate; it also must be presented in a way that common users, who may not have any experience with health or medicine, are able to understand.

Our study’s results highlight a crucial development in the communication of health information: Instagram, a platform more commonly associated with social networking and entertainment, is being increasingly used for health-related purposes. The results of 205 participants who acknowledge they use Instagram to get health information are significant for public health communication work. Moving to digital platforms represents a change in the way people access and use health information, and to maintain public safety and confidence, content quality must be evaluated. Although a significant number of participants reported that they were not looking for health information on Instagram, this still generates a debate about the wide range of ways in which people look for health information and the role of social media in public health literacy. It raises the question of why these users refuse to utilize Instagram for health-related purposes: Is it because they lack confidence in the platform? Do they prefer traditional information sources? Or are they just unaware that the platform offers this kind of information? The results of our research about the use of Instagram for health information indicate an overall increase in the digital age: people are using easily available and captivating platforms to get the health information that need. This means that it is crucial to make sure that information on health is accurate backed by solid data, and communicated in a way that the public can understand and use.

### Limitation

This research has some limitations. Self-reported measurements, which depend on recall and social desirability biases, are used in the survey-based information collection process. Furthermore, there is a chance that the convenience sampling method employed did not produce an outcome that is typical of Azerbaijan’s entire population, which would restrict how broadly the results may be applied. Moreover, the statistics may not accurately represent the variety of medical information available on Instagram at present, given the platform’s quickly changing landscape. As a final point, the research did not distinguish between user-generated information and professionally created content, which might differ greatly in accuracy and quality.

### Recommendations

Instagram has emerged as a significant medium for medical professionals in the age of digital health communication. Certain strategies, however, are essential for making sure that the data is both reliable and impactful:

- Medical professionals should make sure that the information they give is supported by current, peer-reviewed studies. Simplifying challenging medical terminology while preserving accuracy can help make evidence more accessible to a larger audience.
- Doctors should include citations or references for the content that they share, if possible. This creates trust and allows followers to explore further if they desire.
- Addressing questions and concerns in the comment area may boost legitimacy and allow for clarifications, reducing the possibility of misinformation spreading.
- Healthcare professionals including influencers may collaborate on live questions and answer sessions, discussions, or webinars. This can help illuminate complicated medical topics while providing expert knowledge to followers immediately.
- When influencers share or recommend components written by healthcare experts, the availability of solid data rises.
- By initiating joint campaigns to raise awareness about health, the influencer’s creativity and the doctor’s experience may be blended to make certain that the point of view is both compelling and correct.

The recommendations that follow are suggested to maintain the validity of health components on Instagram:

- Content must explicitly differentiate between evidence-based knowledge, personal experiences, and perspectives.
- Use authentic images or illustrations for visual ethics. Avoid dramatization and the use of misleading graphics which could misrepresent the content and cause an unintended effect.
- Health regulations and recommendations vary. Update posts on a regular basis to include the most recent findings and withdraw or replace outdated posts.
- Promote peer evaluations of content before posting it. Feedback from others may assist with ensuring accuracy and comprehensiveness.

## CONCLUSION

- 65 % of the respondents indicated they had used Instagram at least once to find health-related resources.
- Subscribers on Instagram utilized the platform to search for health information with various regularity; among those who never, rarely, or sometimes watched health information on Instagram, there was an equal distribution.
- A large percentage of participants agreed that the medical information they got from Instagram was helpful, demonstrating that users of the platform have accepted information about health.

Several fields remain unexplored in the fast-developing area of digital health communications. The effectiveness of Instagram health campaigns is a good example of that. As the platform’s popularity expands, it is vital to understand how the platform’s wider user population interacts with and reacts to health-related material. How might Instagram campaign visuals, stories, and interactive features impact health behavior and perceptions?

Furthermore, the utilization of Instagram influencers in health education generates a curious junction between popular culture and medical communication. Bloggers have a capacity to impact public opinion and behavior due to their large numbers of followers as well as their perceived authenticity. However, the extent to which they share accurate and valuable health information as opposed to possibly misleading their audience or making profitable content remains an issue of concern.

Moreover, a comparison between various social media platforms in the context of health-related information spread could offer crucial insights. How does Instagram’s engagement, reach, and effect on health information compared to websites like Facebook, Twitter, and TikTok? Does the platform’s nature - its fundamental audience, style of content display, and algorithms - influence the efficacy of health communications?

By studying these subjects, academics and practitioners may better personalize health communication techniques, guaranteeing that they draw on the positive aspects of each platform whilst minimizing any potential disadvantages.

## Data Availability

All data produced in the present study are available upon reasonable request to the authors.

## Acknowledgements

The authors would like to acknowledge and thank the students who took the time to participate in the study.

## Availability of data and materials

The datasets generated during and/or analyzed during the current study are available from the corresponding author on reasonable request.

## About the Author(s)

**Bahar Graefen, MD, MScPH, PhD** is Fulbright Scholar and working as Postdoctoral Visiting Researcher at Chicago State University. She is a member of the American Association of Immunologists (AAI), American Society for Microbiology (ASM) and American Public Health Association (APHA).

**Nadeem Fazal, MD, PhD** is Professor of Microbiology & Immunology at Chicago State University. He is a member of the American Association of Immunologists (AAI) and American Society for Microbiology (ASM). His areas of expertise include Immunology, Microbiology, Cell Biology, and Infectious Diseases.

